# ChatGPT in Occupational Medicine: A Comparative Study with Human Experts

**DOI:** 10.1101/2023.05.17.23290055

**Authors:** Martina Padovan, Bianca Cosci, Armando Petillo, Gianluca Nerli, Francesco Porciatti, Sergio Scarinci, Francesco Carlucci, Letizia Dell’Amico, Niccolo’ Meliani, Gabriele Necciari, Vincenzo Carmelo Lucisano, Riccardo Marino, Rudy Foddis, Alessandro Palla

## Abstract

**Objectives:** The objective of this study is to evaluate ChatGPT’s accuracy and reliability in answering complex medical questions related to occupational health and to explore the implications and limitations of AI in occupational health medicine while providing recommendations for future research in this area and informing decision-makers about AI impact in healthcare.

**Methods:** A group of physicians was enlisted to create a dataset of questions and answers on Italian occupational medicine legislation. They were divided into two teams, each assigned to a different subject area. ChatGPT was used to generate answers for each question with/without legislative context. The doctors evaluated *human* and generated answers in blind, with both teams reviewing each other’s work.

**Results:** Occupational physicians perform better than ChatGPT in generating accurate questions on a 5-point likert score, while ChatGPT with access to legislative context is comparable to professional doctors in providing complete answers. Still, we found that users tend to prefer answers generated by humans, indicating that while ChatGPT is useful, users still value the opinions of occupational medicine professionals.

**Conclusions:** The study evaluated ChatGPT’s effectiveness in occupational medicine and identified crucial factors for its responsible use. It emphasizes ongoing dialogue and reflection for AI development in healthcare. ChatGPT provides 24/7 assistance to occupational physicians, increasing efficiency and reducing costs, monitoring workers’ health, and offering personalized service. It has the potential to transform occupational medicine and create safer work environments.

## Introduction

Artificial intelligence (AI) has become an increasingly popular topic in healthcare to develop predictive models for disease diagnosis and treatment, improve patient outcomes, and optimize resource utilization [1]. In radiology, for instance, AI algorithms have been developed to analyze medical images such as computed tomographic (CT) [2] and magnetic resonance imaging (MRI) [3] scans and detect abnormalities that may be missed by human radiologists. They are also used in public health to identify infectious diseases, in clinical practice to retrieve medical data for diagnosis, and in clinical trials to support decision-making for trial design and monitoring outcomes and side effects [4]. AI has also been employed in cardiology to enhance healthcare standards, improve disease surveillance, optimize intervention timing, and for more accurate outcomes’ predictions [5]. AI has numerous potential benefits in healthcare, with new technologies and applications emerging frequently. One such application is the use of AI chatbots like ChatGPT, which can aid healthcare professionals in patient care and support. ChatGPT is a publicly available chatbot created by OpenAI, a non-profit AI research and development company. It was released in November 2022 and can be accessed at chat.openai.com. AI chatbots typically consist of a chat interface and a Large Language Model (LLM) based on the transformer architecture [6].

Using a chatbot involves inputting a prompt in plain, natural language to initiate a” session”. This prompt is typically a query and can be submitted in various human languages.

### KEY LEARNING POINT

<u>What is already known about this subject</u>

- ChatGPT can generate coherent and compelling text, intelligently responding to a wide range of questions and requests.
- ChatGPT has the potential to revolutionize modern medicine by its ability to process vast amounts of data, provide personalized support to patients, enhance communication between doctors and patients, and offer innovative solutions for diagnosis, treatment, and medical research.
- The effectiveness of ChatGPT in answering complex occupational health questions and its potential implications and limitations in occupational medicine are still being explored through research.

<u>What this study adds</u>

- The study offers insights into the performance of ChatGPT when compared to occupational physicians in generating questions and answers related to occupational medicine.
- The study identifies crucial factors for the responsible use of ChatGPT in occupational medicine and emphasizes ongoing dialogue and reflection for AI development in healthcare.
- The study shows that while ChatGPT can provide 24/7 assistance to occupational physicians, increasing efficiency and reducing costs, users still value the opinions of human occupational medicine professionals.

<u>What impact this may have on practice or policy</u>

- The study presents novel possibilities in the realm of occupational medicine, showcasing how the utilization of ChatGPT has the potential to bring about substantial enhancements in promoting health and well-being within work environments.

The chatbot responds with a natural language reply that pertains to the prompt, resulting in an experience like a conversation between two individuals. ChatGPT was trained on a vast dataset of internet data from multiple sources [7] and fine-tuned on instruction-based following and reinforcement learning from human feedback [8]. Prompts can take the form of questions or specific directives, and can incorporate diverse data inputs like research papers, mathematical equations, and spreadsheets [9] [10].

The first potentials applications of ChatGPT are in medical education, scientific research, medical writing and diagnostic decision-making. [11]. One of the advantages of using ChatGPT in medicine may be its potential ability to analyze large amounts of data which may include scientific articles and medical reports, as well as patient records, all of which can provide in the development of clinical decision [12].

ChatGPT is a first-generation AGI (Artificial General Intelligence) with impressive achievements but also has limitations. Its large-scale neural network requires substantial computing power, making it challenging to deploy outside of large tech corporations. The cloud-only nature of the system poses privacy concerns, especially in sensitive applications like healthcare. ChatGPT cannot directly extract information from external sources like medical journals, limiting its accuracy in the medical field. The training data is capped to the end of 2021, preventing it from staying updated with the latest trends and news. Due to these limitations, ChatGPT can generate inconsistent or contradictory responses that do not align with medical guidelines [11]. There are also concerns about the ethical implications to consider regarding conversational AI in medical practice, as well as the potential for bias and errors in the data used to train AI algorithms. Legal implications of using these technologies cannot be downplayed. For instance, determining liability in case of an inevitable mistake by an AI physician is yet to be established [13].

There has been increasing interest in integrating AI research on worker safety, health, and well-being in the work environment [14]. ChatGPT, as one of the most promising applications of AI, has not yet been evaluated in in the international context of occupational medicine.

The EU has set basic safety and health principles that member states, including Italy, must implement. Italy has chosen a single legislative framework for occupational safety and health, contained in Legislative Decree no. 81/2008, also known as “Testo Unico sulla Salute e Sicurezza sul lavoro” [15]. The D.lgs 81/08 covers various topics related to occupational health, including risk assessment, medical surveillance, training, and emergency preparedness. It has been updated with specific directives related to workplace hazards over time, and periodically amended. Trade unions, employers, and professional councils can submit general questions to the Interdepartmental Commission, which evaluates safety and health regulation implementation and proposes legislation. The Commission’s answers serve as interpretive criteria for supervisory activities and implementation of Italian occupational safety and health laws.

This study evaluates ChatGPT’s effectiveness in answering questions related to D.lgs. 81/08 in Italian occupational medicine. Objectives include assessing the accuracy and reliability of ChatGPT in generating answers to complex medical questions, exploring potential implications and limitations of using ChatGPT in occupational health medicine, and providing recommendations for future research. The study aims to offer insights into using AI in healthcare and inform decision-makers about implementing AI technologies in occupational health medicine.

## Methods

The study involved twelve physicians, comprising of resident and specialist physicians in occupational medicine, who were divided into two groups labeled” red” and” blue”. The work was supervised by a full professor of occupational medicine.

The two groups of physicians were given the task of creating questions and their corresponding answers on the primary topics of occupational medicine. Each doctor was assigned a specific topic, and they were instructed to refer to the most recent version of D.lgs 81/08 as the basis for their questions. The topics covered the main hazards present in workplaces, including safety, biological, physical, ergonomic, chemical, and work organization hazards. In particular the selected topics are: workplaces and use of work equipment, manual handling of loads, use of video terminal, physical hazards, chemical hazard, asbestos, carcinogenic and mutagenic agents, biological and explosive atmospheres in the workplace. They were chosen to cover a wide range of the legislative context. Also, the physicians were instructed to create questions related to a specific chapter of D.lgs 81/08, known as “interpelli”. As stated in the Introduction, the Interdepartmental Commission is responsible for addressing issues related to D.lgs 81/08. The answers to the” interpelli” questions serve as interpretative and guiding criteria for the implementation of the regulations outlined in the latest published version of D.lgs 81/08. Each group member was required to input their assigned questions and answers on a dedicated Google Form platform, with their group (red or blue) specified beforehand.

We used OpenAI ChatGPT API to reply to the physicians’ questions. The method employed two distinct approaches. In the first approach, the Vanilla ChatGPT, the questions generated by the two groups were fed directly into ChatGPT. To mimic a real use-case scenario both questions and answers were submitted in Italian. ChatGPT system message serves as the primary instructions for the model, and it can be tailored to include various information in the system role. This can comprise a brief overview of the assistant, its personality traits, specific instructions, or guidelines that you want the assistant to adhere to, or relevant data or information that the model should have, such as frequently asked questions. It is possible to personalize the system role to suit specific use cases.

Although the system role/message is not mandatory, we found out that high quality system messages improve the overall quality of the generated replies.

In the second approach, ChatGPT + Context, the reference legislative context from the D.lgs 81/08 was also passed alongside the questions. To retrieve the context, we used semantic search with embeddings. Text embeddings refer to a technique used in natural language processing (NLP) and machine learning, which involves the conversion of sentences into numerical vectors. In simpler terms, text embeddings are a way of representing words and paragraphs as numbers, which can be more easily analyzed and processed by computers. We used text embeddings to perform semantic search to retrieve the relevant document from the D.lgs 81/08 as the latter is a type of search that goes beyond traditional keyword-based search and tries to understand the meaning of the query and the content being searched.

The evaluation of generated responses involved a qualitative assessment of the accuracy, precision, completeness, usability, and relevance of each question and its three generated response options. Each question with its three response options was evaluated by each physician who was blinded to the group that had generated the question. The evaluation criteria used to assess the quality of the generated responses are briefly described as follows:

- Accuracy: was the answer correct and aligned with the actual answer.
- Precision: was the answer concise and to the point, without being overly verbose or ambiguous.
- Completeness: was the answer covering all aspects of the question.
- Usability: was the answer easy to understand and use by.
- Relevance: was the answer appropriate and relevant to the question asked.

Additionally, a Likert 5-point scale was used to measure the absolute value of accuracy and completeness of each generated response. This scale allowed for a more fine-grained evaluation of the quality of the responses, by providing a numerical value to indicate the extent to which the response was accurate and complete. The Likert scale ranged from 1 (not accurate/complete at all) to 5 (completely accurate/complete), with intermediate values indicating varying degrees of accuracy and completeness.

This approach allowed for a more objective and quantitative evaluation of the quality of the generated responses, in addition to the qualitative evaluation based on the criteria of accuracy, precision, usability, and relevance. The use of both qualitative and quantitative evaluation methods provided a comprehensive assessment of the quality of the generated responses and allowed for a more robust analysis of the results.

To facilitate the evaluation process, a website was created where each physician could login and access the questions and response options for evaluation. The website provided a user-friendly interface that allowed the doctors to evaluate the responses based on the predefined criteria and using the Likert 5-point scale. The layout of the website was carefully crafted to ensure that the questions and their corresponding three options were always visible to the users. This approach was implemented to simplify the decision-making process for the evaluators.

To ensure that the evaluations were as comprehensive and unbiased as possible, both groups of doctors were given the opportunity to review the questions and answers of the other group and the order of each evaluation was randomized between different users. This approach was implemented to minimize potential cross-talks between evaluations and ensure that all questions were covered as thoroughly as possible. For the same reason, the answers order (users’, ChatGPT, ChatGPT + Context) is randomized in any evaluation to avoid any implicit bias. The website was designed in such a way that participants could only proceed to the next evaluation after completing all fields.

This feature ensured that all evaluations were completed in full and that there were no missing data. Following the completion of the evaluation phase, a questionnaire survey was conducted to investigate the specific errors encountered in the responses generated by ChatGPT + Context. The questionnaire was administered alongside the poorly rated questions, which scored 2 or less on the Likert accuracy scale, to gain insights into the nature of these errors.

## Results

Figures 1 shows the relative likert score of ChatGPT, ChatGPT + Context and user generated answer. In terms of accuracy the users’ answers have been generally evaluated higher, followed by the ChatGPT + Context and the vanilla ChatGPT ones.

**Figure 1:**
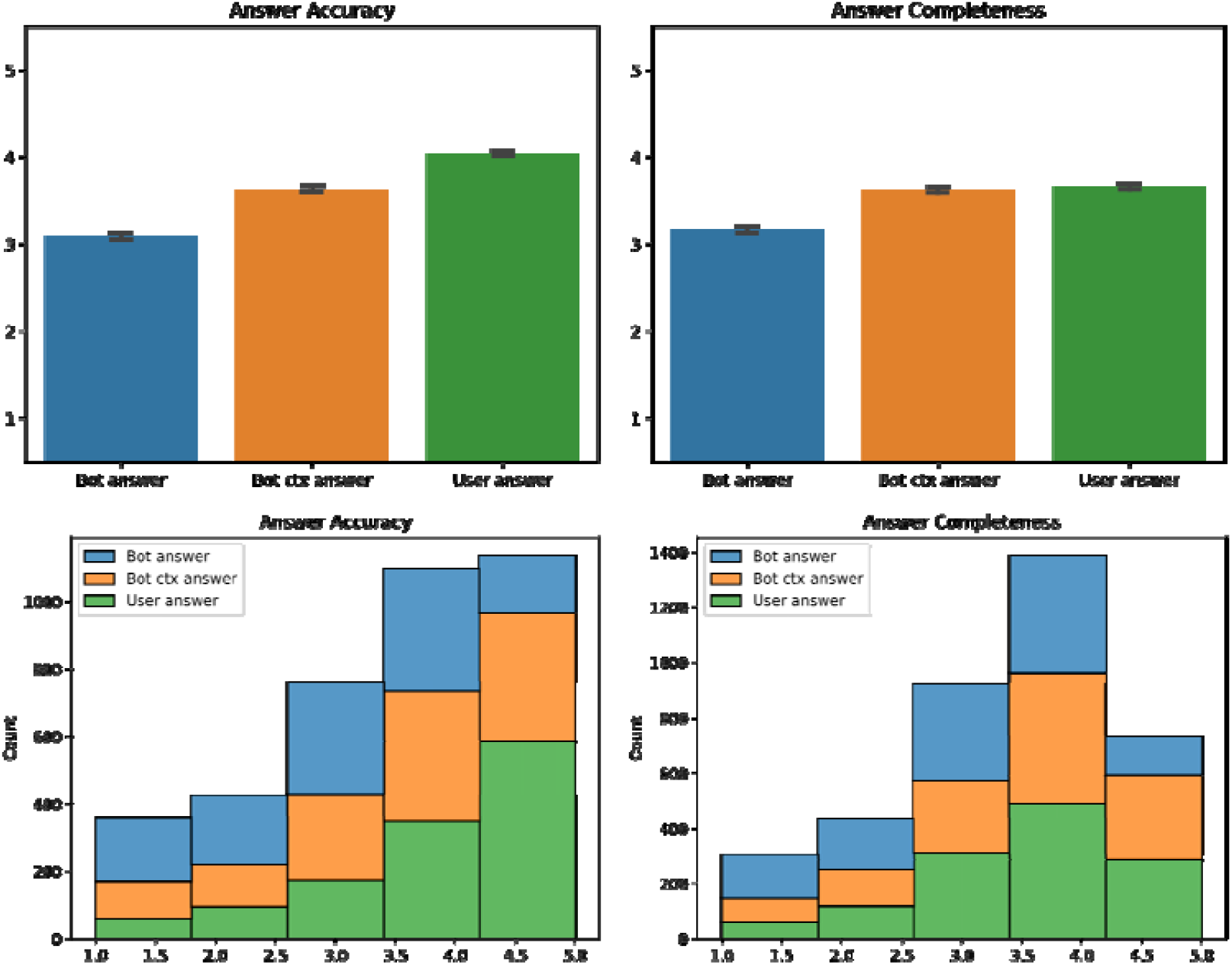
Accuracy and Completeness likert score and distribution for ChatGPT, ChatGPT + Context and Users’ generated question

The accuracy scores for the responses generated by the physicians were significantly higher than those generated by the ChatGPT model, with an average score of 4. 042 ± 0.032 for the physicians, 3.361 ± 0.035 for ChatGPT + Context and 3.091 ± 0. 035 for the vanilla ChatGPT model. The difference was statistically significant with a p-value less than 0. 001 in all cases.

The completeness scores for the responses generated by the physicians were significantly higher than those generated by the ChatGPT model, with an average score of 3.658 ± 0. 030 for the physicians, and 3.159±0.033 for the ChatGPT model. The difference was statistically significant with a p-value lower than 0. 001. However, the difference in completeness scores between responses generated by the physicians and ChatGPT + Context model was not statistically significant, with a p-value of 0.392, meaning that physicians and ChatGPT + Context tend to generate answers that are evaluated in the same ways in terms of completeness.

All results are summarized in Table 1.

**Table 1:**
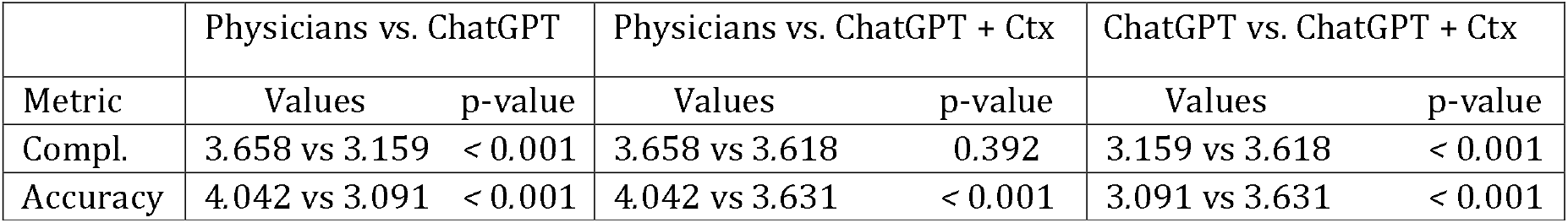
Comparative scores and p-values for Physicians’, ChatGPT and ChatGPT + Context

Figure 2 shows that the completeness of the responses generated by the ChatGPT + Context model was not significantly affected by the utilization of the full D.gls 81/08 legislative context instead of only a part of it. The average completeness score for the full context was 3.618, while the partial context scored the same at 3.618, with a p-value of 0.998. However, the accuracy of the responses generated by the ChatGPT + Context model was significantly affected by the presence of the full context. The average accuracy score for the full context was 3.631, while the partial context scored lower at 3.382, with a p-value of 0.017.

**Figure 2:**
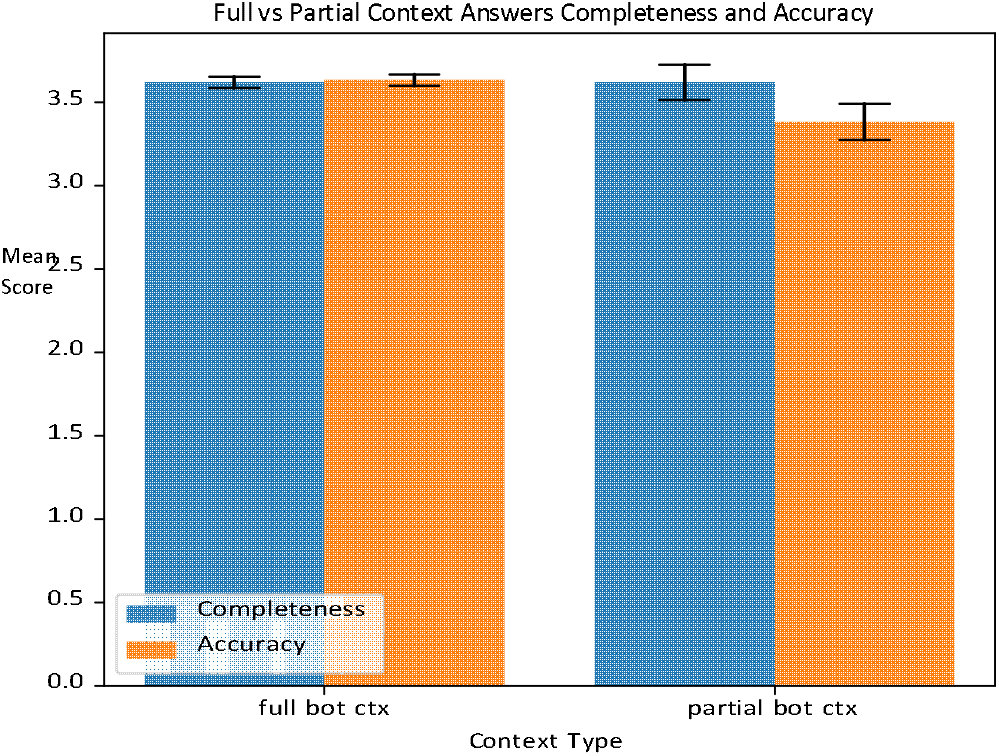
Accuracy and Completeness likert for Full vs Partial Context generated question

Overall, the results suggest that while the ChatGPT + Context model can generate responses that are reasonably complete and accurate, there is still room for improvement to match the quality of responses generated by human experts. Additionally, the use of full context drastically improves the accuracy of the model’s responses, confirming semantic search as a key factor in the overall response quality.

The results of the evaluation showed that ChatGPT + Context responses were generally preferred instead of the vanilla model ones in all five metrics. However, users were found to have a stronger preference for human generated responses in every metric, indicating that there is still room for improvement in the generated responses. Results are shown in Figure 3.

**Figure 3:**
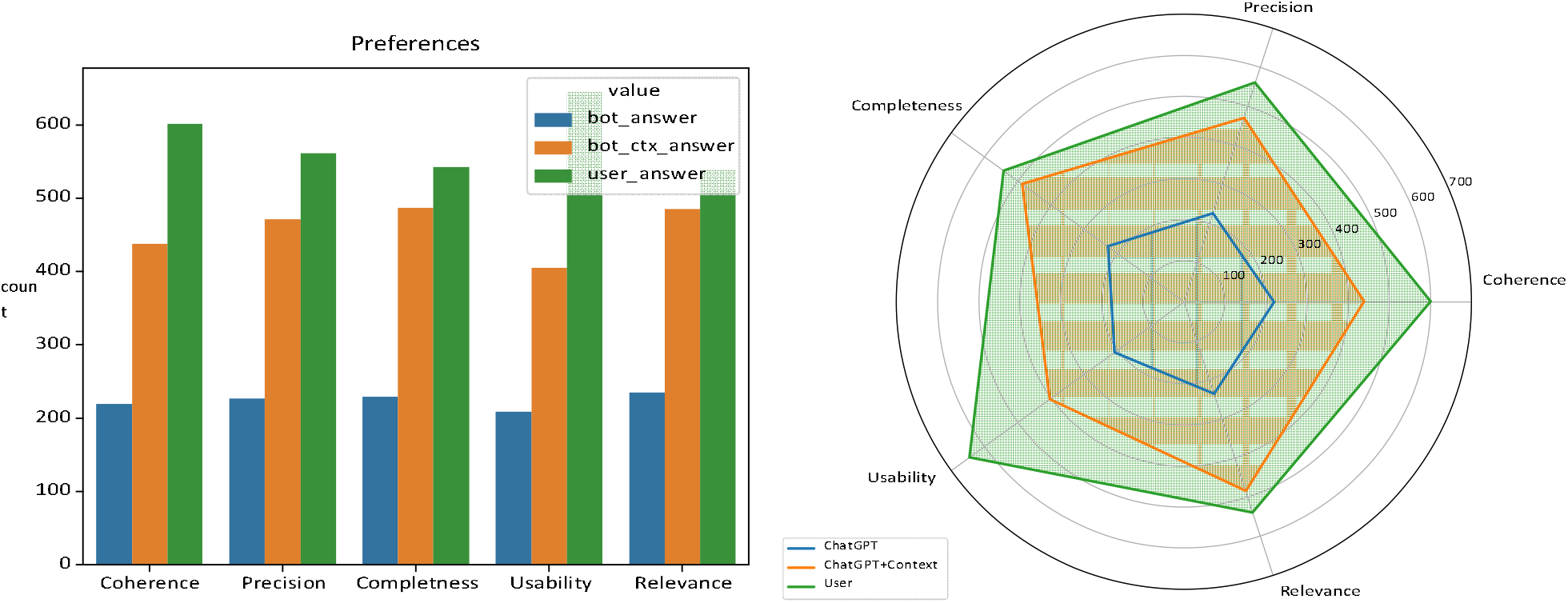
User’s preferences

Regarding the metric of Accuracy, the ChatGPT model had 220 preferences, ChatGPT + Context 438 preferences and users were found to be more accurate, with 601 preferences. Similarly, the Precision metric showed ChatGPT with 227 preference, ChatGPT + Context with 471 preferences, and users with 561 preferences.

The Completeness metric showed a similar pattern, with ChatGPT having 229 preferences, ChatGPT + Context having 487 preferences and users having 543preferences. The Usability metric showed ChatGPT with 209 preferences, ChatGPT + Context with 405 preferences, and users with 645 preferences.

Finally, the Relevance metric showed ChatGPT with 235 preferences, ChatGPT + Context with 485 preferences, and users with 539 preferences. Overall, the results suggest that while ChatGPT + Context performed reasonably well, there is still room for improvement in generating responses that are as accurate, precise, complete, usable, and relevant as human-generated ones.

Following the evaluation, a questionnaire was dispatched to users to determine the nature of errors in ChatGPT+Context responses. The questionnaire was sent together with the poorly rated questions (2 or less on the Likert accuracy scale). The results are depicted in Figure 4. The most common cause of error was found to be incorrect content, followed by improper context. Upon re-analyzing the generated responses, it was observed that, in most cases, the error originated from an incorrect reference context provided to ChatGPT by the semantic search process. This suggests that to enhance the system further, significant efforts should be made to minimize errors in context semantic search and provide more contextual information to the AI. We also found that ChatGPT quality degrades if the user provides a question with roman numbers or acronyms.

**Figure 4:**
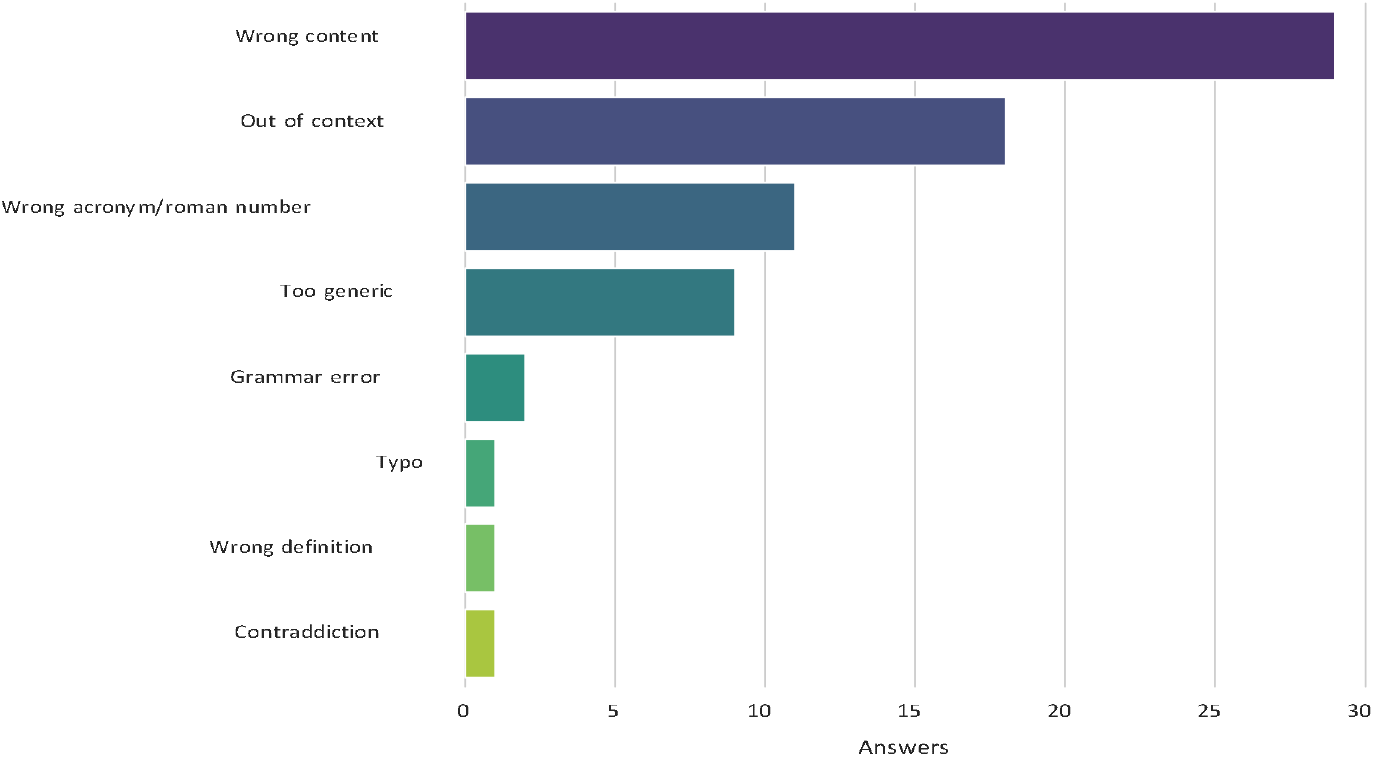
Sources of errors in ChatGPT+Context answers

## Conclusion

The evaluations conducted in this study provide valuable insights into the performance of ChatGPT in comparison to occupational health physicians in generating questions and answers related to occupational health. While ChatGPT may not outperform physicians in generating accurate questions, it is still a valuable tool, especially when equipped with legislative context, as it can provide complete and informative answers to users.

Moreover, users tend to prefer user-generated answers, indicating the importance of integrating user participation in the development of such tools. Overall, this study highlights the potential of ChatGPT as a useful tool in the field of occupational health, but also underscores the importance of considering the limitations of artificial intelligence and the value of human expertise in this field. If these limitations could be overcome, the chatbot could be a useful tool to help the occupational physician with the more mnemonic and notional aspects, leaving the task of making decisions based on ethics, guidelines and of professional experience to the *human*.

ChatGPT’s has ethical, legal, and societal implications. Data privacy and bias, liability, and responsibility are among the concerns to consider, with a need for transparency, accountability, and safeguards. Its impact on health and safety, as well as societal values, requires critical reflection. Therefore, a responsible, ethical, and reflective approach is necessary for their development and use. It is important for developers, practitioners, and policymakers to work together to ensure that the use of such tools is ethical, transparent, and accountable, and that appropriate safeguards are in place to protect both users and professionals in the field of occupational health.

ChatGPT has the potential to transform the landscape of occupational medicine. The use of ChatGPT and similar AI can bring numerous benefits to the field, including medical assistance, increased efficiency, cost reduction, health monitoring, and personalized services. One of the major advantages of ChatGPT is the ability to provide round-the-clock assistance to occupational physicians. With ChatGPT, occupational physicians can ask questions about workplace safety, workers’ rights, safety protocols, and more at any time, and receive immediate responses. ChatGPT can provide efficient service by responding instantly to their queries about the current regulations concerning the protection of health and safety at work.

Moreover, ChatGPT can be used to monitor worker health and provide guidance on healthy lifestyle choices and prevention of work-related illnesses. This can help to promote a healthy and safe work environment and ensure that workers are aware of potential health risks in the workplace.

Based on the findings of this study, several recommendations can be made for the use of ChatGPT in occupational health medicine. Firstly, it is important to ensure that ChatGPT is trained on diverse and representative data to mitigate potential biases in its responses. Additionally, ChatGPT should be used as a tool to support human expertise, rather than replace it, and should not be relied upon as the sole source of information. It is also recommended that users of ChatGPT have access to legislative context to ensure that responses are complete and accurate. Furthermore, it is important to implement appropriate safeguards to protect the privacy and security of user data when using ChatGPT. Finally, ongoing critical reflection and dialogue are necessary to ensure that the use of ChatGPT in occupational health medicine is responsible, transparent, and accountable, and that it ultimately serves the best interests of individuals and society.

## Data Availability

All data produced in the present study are available upon reasonable request to the authors

